# Altered amygdalar emotion space in borderline personality disorder normalizes following dialectical behavioral therapy

**DOI:** 10.1101/2023.01.14.23284531

**Authors:** Seth M Levine, Katharina Merz, Daniel Keeser, Julia I Kunz, Barbara B Barton, Matthias A Reinhard, Andrea Jobst, Frank Padberg, Corinne Neukel, Sabine C Herpertz, Katja Bertsch, Richard Musil

**Author notes:** **Correspondence** Dr. Seth Levine, Division of Clinical Psychology and Psychotherapy, Department of Psychology, LMU Munich, 80802 Munich, Germany. Shared first authorship (Seth M Levine, Katharina Merz). Shared senior authorship (Katja Bertsch, Richard Musil).

## Abstract

Borderline personality disorder (BPD) is a mental health condition characterized by an inability to regulate one’s emotions or accurately process the emotional states of others. Previous neuroimaging studies using classical univariate analyses have tied such emotion dysregulation to aberrant activity levels in the amygdala of patients with BPD. However, multivariate analyses have not yet been utilized to investigate how representational spaces of emotion information may be systematically altered in patients with BPD. To this end, patients performed an emotional face matching task in the MR scanner, before and after a 10-week inpatient program of dialectical behavioral therapy (DBT). Representational similarity analysis of the amygdala revealed a negative bias in the underlying affective space (in that activity patterns evoked by angry, fearful, and neutral faces were more similar to each other than to patterns evoked by surprised faces), which normalized after DBT. This bias-to-normalization effect was present neither in patients’ objective-selective cortex nor in amygdalar activity patterns of a group of healthy volunteers. Such findings suggest a more refined role for the amygdala in the pathological processing of perceived emotions and may provide new diagnostic and prognostic imaging-based markers of emotion dysregulation and personality disorders.

## 1 INTRODUCTION

Borderline personality disorder (BPD) is a severe mental disorder affecting approximately 1.7% of the population and 15 – 28% of inpatients [1]. It is characterized by a pattern of instability in affect, self-image, and interpersonal relations, as well as impulsivity, risk-taking behavior, and hostility [2]. According to a prominent theory [3], emotion dysregulation is conceptualized as the core feature of BPD, rendering it a primary target for evidence-based interventions, such as Dialectical Behavioral Therapy (DBT). Emotion dysregulation comprises faster and elevated responses to stimuli, slower return to baseline, and fewer adaptive (and more maladaptive) regulation strategies [4]. In DBT, patients are trained to, for example, better differentiate their own emotions and decide which emotions are adaptive (and which are overbearing) [3]. This form of therapy has been shown to have a mitigating effect on emotion dysregulation [5].

Previous investigations into emotional processing of patients with BPD have shown heightened emotional sensitivity [6], negativity biases [7, 8], and altered processing of facial expressions compared to healthy individuals [9, 10]. At the neurobiological level, univariate analyses of functional neuroimaging data from patients with BPD have consistently implicated aberrant activity levels in the amygdala in altered emotional processing [11, 12, 10, 13, 14], whereas a normalization of such amygdalar activity has been reported following DBT [15, 16, 17]. Such neuroimaging findings have supported theories designating the amygdala as a key brain region in emotion regulation [18, 19].

However, univariate analyses of functional neuroimaging data have found limited success in generating reliable biomarkers of mental disorders [20]. To that end, the adoption of multivariate methods from cognitive neuroscience has attempted to close the explanatory gap between biological psychiatry and neuroscience. Representational similarity analysis (RSA) [21], a form of multivariate pattern analysis [22] that allows researchers to understand the relative informational content represented by multivariate activity patterns, has only recently been utilized to examine how the cognitive structure of information is altered in different patient groups, such as individuals with post-traumatic stress disorder [23], autism [24], and schizophrenia [25]. So far, no study has employed RSA to investigate high-dimensional neural emotion spaces [26, 27, 28] in individuals with BPD.

As such, the present study sought to extend prior neuroimaging findings by using RSA to explore whether neural emotion spaces, measured using a classic perceptual matching task of emotional facial expressions [29], show systematic alterations [30] before and after BPD patients underwent a 10-week program of DBT.

## 2 METHODS

### 2.1 Participants

Twenty-one inpatients between the ages of 19 and 54.5 years (mean age [SD] = 27 [±10] years; 12 females, 6 males, 2 transgender males, 1 unspecified) were recruited to this study as part of an ongoing trial for BPD patients and patients with persistent depressive disorder at the Department of Psychiatry and Psychotherapy of the University Hospital LMU Munich [31, 32, 33]. Six participants did not participate in the second neuroimaging session, as they were either discharged early from the clinic or refused to participate in the second session. As such, full datasets for the remaining 15 participants between the ages of 19.8 and 54.5 years (mean age [SD] = 28.6 [±11] years; 8 females, 6 males, 1 transgender male) were included in the present analysis. All experimental procedures were approved by the ethics committee of the Faculty of Medicine of the LMU and comply with the Declaration of Helsinki following its most recent amendments. Participants provided written informed consent before participating in the study.

Regarding common comorbidities of BPD, following a SCID-5-CV [34, 35] assessment at the beginning of the treatment, 12 patients had a current major depressive episode, eight patients were diagnosed with life-time PTSD (current symptomatology, n = 5), five patients showed a life-time binge eating disorder (current symptomatology, n = 1), and one participant showed a life-time (and current) bulimic eating disorder.

### 2.2 Clinical scales

For clinical assessment, we administered the Beck Depression Inventory (BDI-II) [36, 37], the Hamilton Depression Rating Scale (HAMD-24) [38, 39], the Borderline Symptom List (BSL-23) [40, 41], the Borderline Personality Disorder Severity Index Version IV (BPDSI-IV) [42, 43], and the short form of the Childhood Trauma Questionnaire (CTQ) [44, 45] at admission. After the 10-week treatment of dialectical behavioural therapy (DBT) [3], 15 patients completed a second administration of the HAMD-24, 14 completed a second administration of the BPDSI-IV, and 12 completed a second administration of the BDI-II and BSL-23 (Table 1).

**Table 1.** Descriptive statistics for the clinical scores. The (4) and (6) for the BPDSI-IV refer to the subscales for its 4^th^ and 6^th^ symptom areas (i.e., impulsivity and affective instability, respectively). *CTQ scores were only acquired during the first session and therefore do not reflect a difference score. **P-values derive from Wilcoxon signed-rank tests contrasting the median score of the two sessions.

|  | BDI-II | BSL-23 | HAMD-24 | BPDSI-IV | BPDSI-IV (4) | BPDSI-IV (6) | CTQ* |
| --- | --- | --- | --- | --- | --- | --- | --- |
| Median (Post - Pre) | -3.5 | 0.025 | -2 | -5.57 | -0.64 | -0.1 | 48 |
| Interquartile range | (-9, 2) | (-0.52, 0.57) | (-8.5, 4.5) | (-12.39, 1.25) | (-1.275, -0.005) | (-1.7, 1.5) | (34.5, 61.5) |
| P-value** | 0.054 | 0.079 | 0.11 | 0.013 | 0.021 | 0.69 | - |

### 2.3 Experimental paradigm

While lying in the MR scanner, participants performed a classic perceptual matching task [29], in which they were visually presented with alternating blocks of emotional faces (i.e., angry, fearful, neutral, surprised) or shapes (which served as the control condition). On a given trial, participants saw three stimuli (following the emotional theme of the current block) simultaneously: one at the top of the screen (the target stimulus) and two at the bottom of the screen, with the goal of determining which stimulus below is identical to the target stimulus above. Each emotion block lasted 48 seconds, with six stimuli of a given emotional expression appearing in each block for 4 seconds with a variable interstimulus interval of 2 – 6 seconds. Each shape block lasted 36 seconds, with six stimuli of different shapes appearing in each block for 4 seconds with a fixed interstimulus interval of 2 seconds. Inter-block intervals were 12 seconds in duration. The run started and ended with a shape block. The task was administered using Presentation^*®*^ software (Version 18.0, Neurobehavioral Systems, Inc., Berkeley, CA, USA, https://www.neurobs.com/), and stimuli were projected onto a screen that participants viewed using a mirror in the scanner. Responses were provided via an MR compatible keypad (Current Designs, Philadelphia, PA, USA).

### 2.4 Neuroimaging acquisition parameters

Neuroimaging data acquisition was carried out at the Neuroimaging Core Unit Munich (NICUM) of the LMU using a 3T Siemens Magnetom Prisma and a 32-channel head coil (Siemens AG, Erlangen, Germany). Functional sequences consisted of 650 volumes acquired with a T2*-weighted EPI sequence (72 slices per volume in ascending interleaved order with multiband factor 8, voxel size = 2 mm^3^ isotropic, TR = 800 ms, TE = 37 ms, flip angle = 52^0^, FoV = 208 mm). The first five volumes of functional scans were dummy volumes to account for T1-saturation and were discarded prior to image preprocessing. To coregister the functional images with the high-resolution anatomical images, 208 slices of T1-weighted scans were acquired using a magnetization prepared rapid gradient-echo (MP-RAGE) sequence (voxel size = 0.8 mm^3^ isotropic, TR = 2500 ms, TE = 2.22 ms, flip angle = 8^0^, FoV = 256 mm).

### 2.5 Neuroimaging data analysis

Neuroimaging data were analyzed using SPM12, MATLAB R2020a (The Mathworks, Natick, MA, USA), and CoS-MoMVPA [46]. Preprocessing of the neuroimaging data made use of default settings of the SPM12 preprocessing pipeline (https://www.fil.ion.ucl.ac.uk/spm/software/spm12/) and included (1) spatially realigning the functional images to the mean image in the time series using a six-parameter rigid body transformation and 4^th^ degree b-spline interpolation, (2) coregistering the functional images to a given participant’s T1-weighted structural scan, (3) normalizing the coregistered images to a standard 2 mm MNI template using 4^th^ degree b-spline interpolation, and (4) spatially smoothing the images with a Gaussian kernel (8 mm FWHM). Slice-timing correction was not performed, as the task was a block-design.

The preprocessed functional images were analyzed using the general linear model containing one regressor per condition. Regressors corresponding to the task blocks were modeled as box-car functions and convolved with a canonical hemodynamic response function. Motion correction parameters were modeled as regressors of non-interest in addition to a constant term.

### 2.6 Region-of-interest definition

As this paradigm is known to activate the amygdala when contrasting face blocks with shape blocks [29], we sought to determine whether the amygdala also systematically represents *patterns* of multivariate activity pertaining to emotion information; as such, we obtained a bilateral amygdala region-of-interest (ROI) from the probabilistic Harvard-Oxford atlas [47, 48], which was masked at a probability threshold of 0.8, yielding a region size of 209 voxels. Each participant’s whole-brain t-scores (from the parameter estimates generated via the GLM) for the contrasts of interest (i.e., [Anger > Shapes], [Fear > Shapes], [Neutral > Shapes], and [Surprise > Shapes]) were masked using this amygdala ROI for the RSA.

### 2.7 Representational similarity analysis

Per participant, the t-scores within the extracted voxels were Pearson correlated across conditions from the first timepoint and again, separately, from the second timepoint. This procedure yielded six correlation values (i.e.,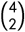) per timepoint, which were visualized as correlation matrices (Fig. 1A–C). The pattern visible in the correlation matrices (Fig. 1A) regarding the difference between the surprise stimuli and the other stimuli (in the first session) led us to investigate between-condition correlations *within* time points, which then allowed us to compare these relative differences *between* time points. To this end, for a given subject and a given time point, we separated correlation values that involved the surprised condition from correlations that did not involve the surprised condition and averaged these two sets independently (i.e., correlations between anger-fear, anger-neutral, and fear-neutral were averaged together, and correlations between anger-surprise, fear-surprise, and neutral-surprise were averaged together). This procedure yielded an “ other vs. surprise” analysis that we investigated before and after therapy (Fisher transforming all participants’ averaged correlation values) via a two-factorial repeated-measures analysis of variance (ANOVA) using SPSS (Version 28.0. Armonk, NY: IBM Corp). Statistical thresholds were set at an *α* level of 5%.

**Figure 1.**
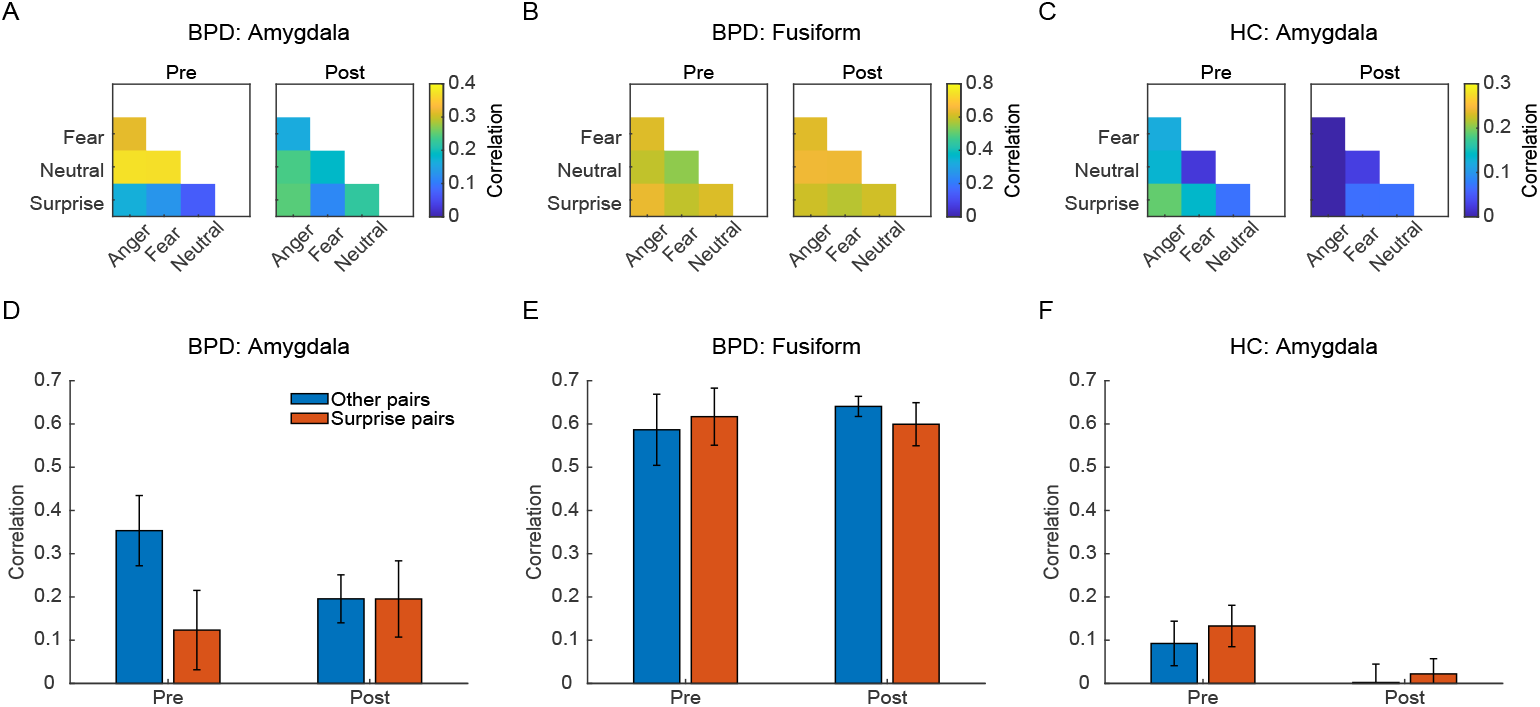
Results from the representational similarity analysis depicted as correlation matrices of the multivariate patterns evoked by the emotional facial expressions for both sessions in **(A)** the amygdala of the BPD patients, **(B)** the fusiform gyrus of the BPD patients, and **(C)** the amygdala of the healthy controls (HC). The amygdalar emotion space of the BPD patients revealed **(D)** a higher degree of similarity between angry, fearful, and neutral expressions (blue bars)—compared to the similarity of surprised expressions with the other facial expressions (red bars)—which normalized in the second session, following DBT (F_1,14_ = 5.027, p = 0.042). This interaction effect from **(D)** was observed neither in **(E)** the fusiform gyrus of patients (F_1,14_ = 0.174, p = 0.683) nor in **(F)** the amygdala of healthy controls (F_1,24_ = 0.63, p = 0.804). Error bars represent SEM.

### 2.8 Regional control

To determine whether the multivariate findings were specific to the amygdala, we also extracted a bilateral ROI from ventrotemporal cortex, which is known to encode object categories [49, 50, 51, 52]. The expectation was that this region would encode face information (and also potentially the corresponding emotional expressions) but that there would be no systematic changes in the representational space following DBT. Specifically, we obtained a bilateral temporo-occipital fusiform ROI from the probabilistic Harvard-Oxford atlas, which was masked at a probability threshold of 0.63 (yielding a region size of 217 voxels), so that the control ROI contained roughly the same number of voxels as the amygdala ROI. Here, we employed a three-factorial (Region X Emotion X Time) repeated-measures ANOVA to compare the results from the amygdala with those from the fusiform.

### 2.9 Healthy volunteer group

An additional follow-up idea sought to determine whether this amygdala-specific effect was also specific to patients. To this end, we incorporated a neuroimaging dataset from 25 healthy individuals (mean age [SD] = 30.2 [±7.8] years; 18 females, 9 males) who underwent the same emotion task in two separate sessions separated by a period of approximately seven weeks [17]. These neuroimaging data were acquired at Heidelberg University between 2016-2018 using a 3T Siemens Tim Trio scanner (Siemens AG, Erlangen, Germany) and a 32-channel head coil. Functional sequences consisted of 40 transverse slices per volume acquired with a T2*-weighted gradient EPI sequence (voxel size = 2.3 mm^3^ isotropic, TR = 2340 ms, TE = 26 ms, flip angle = 80^0^ ; FoV = 220 mm). To coregister the functional images with the high-resolution anatomical images, structural scans were acquired using a T1-weighted MP-RAGE sequence (voxel size = 1 mm^3^ isotropic, TR = 1900 ms, TE = 2.52 ms, flip angle = 9^0^, FOV = 256 mm). See [17], for further details of this dataset.

The same ROI analysis on the amygdala was carried out in the healthy controls, and a three-factorial (Group X Emotion X Time) mixed ANOVA was employed to compare the results from the BPD patients with those from the healthy volunteers.

### 2.10 Follow-up correlation analysis with clinical scales

Given the findings from the similarity analysis, we wanted to explore whether there was any correspondence between the clinical scales and the changes in the amygdalar emotion space. To this end, the difference scores (post minus pre) for each clinical scale (with the exception of the CTQ, which was only administered once) were rank-correlated (using Kendall’s T b) with the interaction values from the activity patterns in the amygdala. In additional to the overall score of the BPDSI-IV, we also used the scores from the subscales for impulsivity (symptom area 4) and affective instability (symptom area 6), as these aspects of BPD have been tied to functionality of the amygdala [53, 54]. Corresponding p-values were generated following 10000 iterations of permutation testing.

## 3 RESULTS

### 3.1 Negative-shifted emotion space in amygdala that normalizes after therapy

Representational geometry of patients’ emotion spaces within the amygdala (Fig. 1A) showed a negative bias in the first session that was not detected in the second session (Time X Emotion: F_1,14_ = 5.027, p = 0.042; Fig. 1D). Specifically, before DBT, activity patterns evoked by angry, fearful, and neutral facial expressions showed a greater average similarity to each other (i.e., “ other pairs”) than to facial expressions depicting surprise (t_14_ = 2.805, p = 0.014). Following DBT, this “ imbalance” in the emotion space was no longer evident, as the representational geometry revealed a more uniform degree of similarity among the activity patterns (t_14_ = 0.005, p = 0.996).

### 3.2 Emotion space in object-selective cortex remains relatively stable

To determine whether the systematic change in the emotion space was specific to the amygdala, we ran the same analysis in the temporo-occipital fusiform, knowing that face information is reportedly encoded by the ventrotemporal cortex. Here the representational geometry showed a dramatically higher *overall* degree of similarity between all facial expressions compared to that of the amygdala (Region: F_1,14_ = 29.995, p = 8.2 × 10^−5^; Fig. 1B); the interaction between emotions and time, as observed in the amygdala, also differed between regions (Region X Emotion X Time: F_1,14_ = 5.866, p = 0.03), with no detectable evidence for such an interaction effect in the fusiform (Emotion X Time: F_1,14_ = 0.174, p = 0.683; Fig. 1E).

### 3.3 Emotion spaces differ between patients and healthy volunteers

The last follow-up control analysis sought to determine whether the dynamic aspect of the emotion space underlying the amygdala was specific to patients with BPD, or whether time alone could explain this effect, in that a similar systematicity would be observable in healthy volunteers at two different points in time. To this end, we applied the same analysis to amygdalar voxels of healthy volunteers, which revealed, firstly, an overall difference between the groups, in that pattern correlations of the patients with BPD tended to be higher than those of healthy volunteers (Group: F_1,38_ = 7.054, p = 0.011; Fig. 1C), but, more importantly, the previously reported interaction effect differed between the groups (Group X Emotion X Time: F_1,38_ = 5.184, p = 0.029) and was not observed in the healthy volunteers (Emotion X Time: F_1,24_ = 0.63, p = 0.804; Fig. 1F). Additionally, the emotion spaces of healthy volunteers did not show any systematic changes in terms of emotions (F_1,24_ = 0.622, p = 0.438) or time (F_1,24_ = 3.685, p = 0.067).

### 3.4 Correlation with clinical scales

Rank-correlating the interaction values from the amygdalar activity patterns with the difference scores in the clinical scales revealed a slight positive correlation between the 4^th^ symptom area of the BPDSI-IV (i.e., “ impulsivity”). Namely, decreasing pattern similarity of the non-surprised facial expressions (with respect to the changing pattern similarity of the surprised facial expressions, i.e., the interaction effect) corresponded to decreasing impulsivity scores (T = 0.35, p = 0.03; Fig. 2). The remaining correlations for the BDI-II (τ = -0.11, p = 0.66), BSL-23 (τ = -0.09, p = 0.64), HAMD (τ = 0.12, p = 0.26), BPDSI-IV: total (τ = 0.05, p = 0.38), BPDSI-IV: “ affective instability” symptom area (T = 0.01, p = 0.46), and CTQ (τ = -0.03, p = 0.56) did not surpass the statistical threshold. All p-values presented here were generated through permutation testing and reflect one-sided statistical tests. None of these correlations survived a correction for multiple comparisons; as such, these results should be considered exploratory.

**Figure 2.**
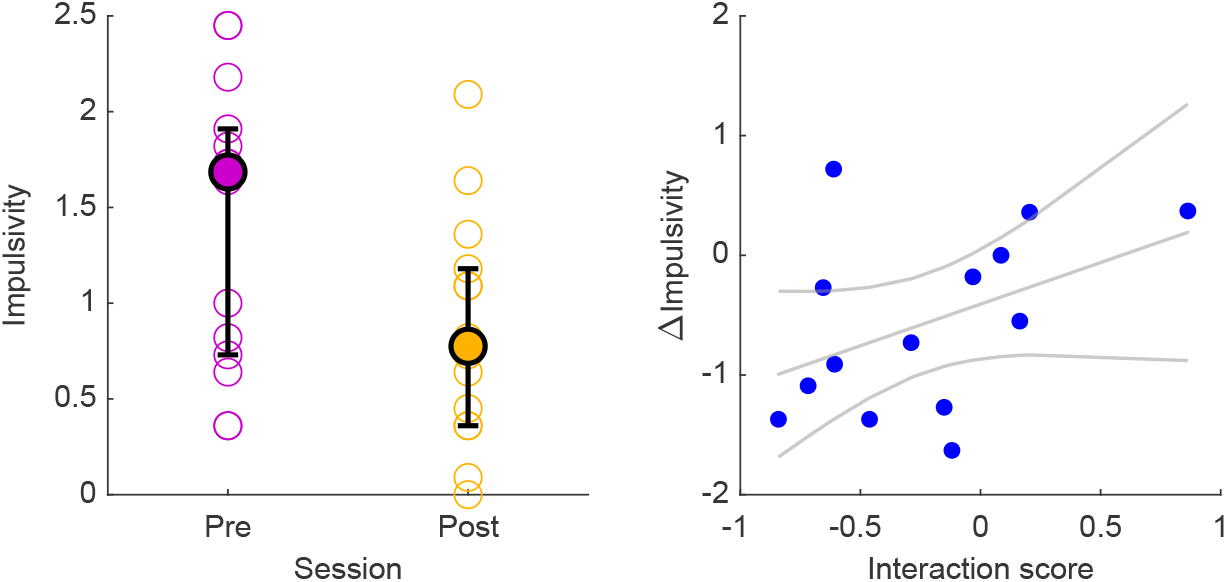
**(Left)** Changes in the impulsivity scores from the BPDSI-IV interview (median and interquartile range overlaid) and **(right)** their respective correlation (Kendall’s T = 0.35, p = 0.03) with the interaction in the multivariate pattern changes (reflecting alterations within the emotion space) in the amygdala. The line of best fit and its corresponding 95% confidence intervals are depicted in gray.

## 4 DISCUSSION

Emotion dysregulation is a core symptom of BPD [55], and DBT focuses on this dysregulation by training patients to differentiate their emotions [3]. Functional neuroimaging studies of such emotion dysregulation in patients with BPD have used univariate analyses to consistently reveal altered activation levels of the amygdala in patients with BPD [56, 57], also with respect to treatment programs incorporating DBT [58]. However, the use of multivariate pattern analysis opens up new avenues for interpreting the role of the amygdala in BPD, as representational similarity analysis (RSA) allows one to hypothesize not only about the *involvement* of a brain region but more specifically about the *representational content* underlying its activity patterns [59].

As such, this study sought to provide a first look into high-dimensional neural representations of perceived emotions in patients with BPD. To this end, we combined RSA with functional MRI to investigate how the representational geometry of emotion information in the amygdala differs in BPD patients before and after DBT. Our findings revealed that, prior to therapy, the representational space of perceived emotions was unusually negative-shifted in patients with BPD, in that angry, fearful, and neutral faces were represented more similarly to each other, while surprised faces were represented less similarly to all other emotions. After therapy, this systematicity normalized, such that all representations of emotional expressions maintained a comparable degree of similarity to each other (i.e., the emotions were more evenly distributed across the representational space). This unexpected structure in the affective representational space of the amygdala is consistent with negativity biases observed in patients with BPD [60] and was detected neither in object-selective (i.e. ventrotemporal) cortex of patients with BPD nor in the amygdala of healthy volunteers.

These findings are supported by prior studies showing that multivariate patterns in the amygdala reflect aversive learning [61], subjective valence [62], and facial expressions [63]. Here, we extend such work by demonstrating that a diagnosis of BPD can also contribute to alterations in amygdalar affective spaces. The specificity of this finding in the amygdala, with respective to the fusiform gyrus, is also corroborated by previous work showing that changes in representational spaces following fear-conditioning occurred in downstream regions involved in affective processing rather than in object-selective cortex [64, 65, 66].

Additionally, Puccetti and colleagues recently employed RSA to demonstrate that a decreased persistence of the amygdala to represent negative information corresponded to higher psychological well-being [67]. This discovery is in line with our result that the amygdalar affective space normalized in patients with BPD following DBT, which raises the question of whether systematic variations in this space might be indicative of meaningful individual differences and have prognostic value. As such, the findings we present here offer a new perspective on the involvement of the amygdala in (pathologically) representing emotion information and may reflect a neural mechanism of emotion dysregulation that classically characterizes BPD.

## 5 LIMITATIONS AND FUTURE OUTLOOK

One of the primary methodological limitations of our study derives from the sample of healthy volunteers having not been specifically matched to the demographics of the BPD patients in the current study and having been acquired on a different MR scanner with different scanning parameters [17]. Therefore, we acknowledge that a more rigorous control sample with matching acquisition protocols would ultimately be necessary; as such, this control analysis represents only a first step in determining the specificity of the effects reported here.

Another limitation of our study involves the extent to which we can associate the representational geometry in the amygdala to specific pathological aspects of BPD. Although the correlation analysis revealed a possible link between the altered affective space and impulsivity scores, the relationship was not particularly robust (as evidenced by the failure of the correlations to survive a correction for multiple comparisons); however, this null effect could simply be due to our small sample size.

As this study is the first to apply RSA to fMRI data of patients with BPD, follow-up work incoporating similar methodology, larger samples, and additional questionnaires (e.g., the Difficulties in Emotion Regulation Scale [68, 69]) is warranted in order to better characterize the relationship between neural representational spaces, emotion dysregulation, and BPD. One idea would involve carrying out several neuroimaging scans throughout the course of a DBT program in conjunction with a dismantling design [58]. This approach could help to constrain our understanding of the relationship between specific aspects of therapy and changes in the neural representational geometry, potentially revealing how such altered representational spaces map onto pathological behavior in BPD, thereby increasing the prognostic value of functional MRI in the clinic. Another idea would involve applying the same analyses to neuroimaging data from a different patient population—also characterized by issues with interpersonal interactions (e.g., depression)— to determine the diagnostic specificity of altered emotion spaces in the amygdala.

## 6 CONCLUSION

Many studies over the past decades have revealed abnormal activation levels of the amygdala as a potential mechanism underlying the behavior of patients with BPD. In this brief report, we provide a first glimpse into the combination of multivariate pattern analysis with functional MRI data acquired from patients with BPD. Before and after patients underwent a 10-week inpatient program of DBT, we used RSA to explore the informational content of activity patterns in the amygdala evoked from a task involving identification of facial expressions. Our approach revealed a negative-shifted representational space before therapy, in which angry, fearful, and neutral faces were represented unusually similarly to one another, while surprised faces were unusually dissimilar to the other expressions. This bias normalized following therapy. Such findings indicate that RSA can reveal novel insights into the neurobiological underpinnings of information processing in personality disorders, which has the potential to increase the diagnostic and prognostic value of functional neuroimaging for clinical psychology and psychiatry.

## Data Availability

All data produced in the present study are available upon reasonable request to the authors

## ACKNOWLEDGEMENTS

This study is part of a clinical trial within the German Clinical Trials Register (Deutsches Register Klinischer Studien [DRKS]) under the identifier DRKS00019821.

## DATA AVAILABILITY

All data produced in the present study are available upon reasonable request to the authors.

## FUNDING INFORMATION

This project was supported by the FöFoLe program (PI: MAR, grant number 996) and the FöFoLe^PLUS^ program (PI: MAR, grant number 003, Munich Clinician Scientist Program) from the Faculty of Medicine of the LMU Munich, as well as from the Friedrich Baur Stiftung (PI: MAR, grant number 74/18).

The Heidelberg study was part of the Clinical Research Group (Klinische Forschungsgruppe [KFO] 256, spokesperson: Christian Schmahl, deputy spokesperson: Sabine C Herpertz) on the “ Mechanisms of Disturbed Emotion Processing in Borderline Personality Disorder” and was funded by grants from the German Research Foundation (Deutsche Forschungsgemeinschaft [DFG], grant numbers: HE 2660/12-2 and BE 5292/3-2).

## CONFLICT OF INTEREST

FP is a member of the European Scientific Advisory Board of BrainsWay Inc., Jerusalem, Israel and the International Scientific Advisory Board of Sooma, Helsinki, Finland. He has received speaker’s honoraria from Mag&More GmbH, the neuroCare Group, Munich, Germany, and BrainsWay Inc. His lab has received support with equipment from neuroConn GmbH, Ilmenau, Germany, Mag&More GmbH, and Brainsway Inc.

RM has received financial research support from the EU (H2020 No. 754740) and served as PI in clinical trials from Abide Therapeutics, Böhringer-Ingelheim, Emalex Biosciences, Lundbeck GmbH, Nuvelution TS Pharma Inc., Oryzon, Otsuka Pharmaceuticals, and Therapix Biosciences.

The other authors declare no financial conflicts of interest.

## AUTHOR CONTRIBUTIONS

AJ: Supervision of conduct, manuscript revision

BBB: Conceptualization and research design (patients), data acquisition and management (patients), manuscript revision

CN: Data acquisition, preprocessing, and management (healthy controls), manuscript revision

DK: Conceptualization and research design (patients), supervision and quality control of neuroimaging (patients), manuscript revision

FP: Conceptualization and research design (patients), supervision of conduct (as principal investigator), manuscript revision

JIK: Data acquisition and management (patients), manuscript revision

KB: Research design (healthy controls and patients), funding acquisition (healthy controls), interpretation of results, manuscript revision

KM: Data acquisition, management, and analysis (patients), manuscript drafting and revision

MAR: Research design (patients), manuscript revision

RM: Conceptualization (patients), supervision of conduct, manuscript revision

SCH: Data acquisition (healthy controls), funding acquisition (healthy controls), manuscript revision

SML: Data preprocessing (patients), data analysis (healthy controls and patients), interpretation of results, manuscript drafting and revision

## Notes

### Clinical Trial

DRKS00019821

### Funding Statement

This project was supported by the FöFoLe program (PI: MAR, grant number 996) and the FöFoLe^PLUS^ program (PI: MAR, grant number 003, Munich Clinician Scientist Program) from the Faculty of Medicine of the LMU Munich, as well as from the Friedrich Baur Stiftung (PI: MAR, grant number 74/18). The Heidelberg study was part of the Clinical
Research Group (Klinische Forschungsgruppe [KFO] 256, spokesperson: Christian Schmahl, deputy spokesperson: Sabine C Herpertz) on the "Mechanisms of Disturbed Emotion Processing in Borderline Personality Disorder" and was funded by grants from the German Research Foundation (Deutsche Forschungsgemeinschaft [DFG], grant numbers: HE
2660/12-2 and BE 5292/3-2).

### Author Declarations

Ethics committee of the Faculty of Medicine of the LMU Munich gave ethical approval for this work.

